# “*I gained a superpower… is this how normal people feel?*”: A qualitative exploration of dietary perceptions and experiences amongst adults living with Cystic Fibrosis in the modulator era

**DOI:** 10.1101/2025.10.26.25338844

**Authors:** Cian Greaney, Katie Bohan, Sarah Tecklenborg, Ciara Howlett, Karen Cronin, Clodagh Landers, Mary Connolly, Derbhla O’Sullivan, Audrey Tierney, Katie Robinson

**Affiliations:** School of Allied Health, University of Limerick, Limerick, Ireland; Food, Diet and Nutrition Research Group, Health Research Institute, University of Limerick, Limerick, Ireland; Cystic Fibrosis Ireland, Rathmines, Dublin, Ireland; Cork University Hospital, Cork, Ireland; St. Vincent’s University Hospital, Dublin 4, Dublin, Ireland; University Hospital Galway, Galway, Ireland; University Hospital Limerick, Dooradoyle, Limerick, Ireland; Centre for Implementation Research, Health Research Institute, University of Limerick, Limerick, Ireland; Discipline of Food, Nutrition and Dietetics, La Trobe University, Melbourne, Victoria, 3086, Australia; Aging Research Centre, Health Research Institute, University of Limerick, Limerick, Ireland

**Keywords:** Cystic Fibrosis, diet, experiences, enablers, barriers

## Abstract

**Background:** Cystic Fibrosis (CF) care has been transformed by modulator therapies, which have extended survival and reshaped daily health management. While CF-specific nutrition guidelines have shifted to focus not just on energy, but also on diet quality, we still know little about how adults living with CF perceive and experience diet in this new treatment era. To address this, the present study aims to explore in depth, the experiences and perspectives of Irish adults living with CF regarding diet and nutrition in a modulator therapy era.

**Methods:** Adults living with CF were recruited via Cystic Fibrosis Ireland networks and specialist clinics across Ireland. Data were generated through virtual focus groups and semi-structured interviews on Microsoft Teams. Transcripts were managed in NVivo® and examined using Braun and Clarke’s inductive thematic analysis framework.

**Results:** Three focus groups (*n* = 9) and 20 semi-structured interviews were completed. Of the 29 participants (male: 62.1%, mean age: 35.0 ± 10.1), one participant was underweight, and 48.3% were overweight/obese. Five interconnected themes were identified: *Legacy effects of CF historic diet*; *Evolving relationship with food*; *Weight gain and altered body image*; *Reclaiming authority over the body*; *Navigating dietary choices*. Participants described persistent anxieties, shifting habits, and the influence of social, emotional, and practical factors on food practices. While the modulator era has led to more autonomy, adults living with CF desire more holistic nutrition guidance, and voiced novel concerns related to their self-image.

**Conclusion:** This study is the first in-depth exploration of Irish adults living with CF, and shows how historic diets, modulators, and daily contexts shape food experiences. Participants reported shifting relationships with food, body image, and autonomy since the introduction of modulators, underscoring the need for holistic, patient-centred nutrition care that integrates medical guidance with lived experience.

## 1. Introduction

Cystic Fibrosis (CF) is an autosomal recessive disorder resulting from mutations in the CF transmembrane conductance regulator (CFTR) gene, with clinical presentation often manifesting as reoccurring pulmonary exacerbations, higher resting energy expenditure from respiratory effort, and nutrient malabsorption, consequentially leading to poorer lung function, nutritional status, and life expectancy (1). The symptom profile in CF increases energy requirements (2), often achieved through high-fat diets (3), which alongside pancreatic-enzyme replacement therapy, has resulted in significant improvements in survival rates amongst people living with CF (PwCF) (4).

In the last decade widespread implementation of highly effective variant-specific therapies (VSTs) (e.g., CFTR modulator therapies) has resulted in unprecedented health improvements for many people with CF (5, 6). Consequently, mean survival age has increased, with many more PwCF expected to live well into adulthood (7, 8). However, despite these positive changes, growing rates of diet-related chronic diseases (i.e., obesity, cardiovascular disease, type-II diabetes) (9, 10), weight gain and changes in body composition have been reported since the introduction of VSTs (11–14). While weight gain is often beneficial, shifting many adults living with CF from underweight into a normal body mass index (BMI) range, evidence indicates a rising prevalence of overweight and obesity (BMI ≥ 25 kg/m²) in this population, as shown in both retrospective (15) and prospective (16) studies. Furthermore, evidence suggests that much of the weight gain observed with Ivacaftor is driven by increases in fat mass rather than lean mass (17). Simultaneously, diabetes has become an increasingly important clinical concern. A large longitudinal analysis of European CF registry data (*N* = 40,096) demonstrated a clear age-related increase in CF-related diabetes, with prevalence rising from 0.8% in those under 10 years, to 9.7% in ages 10–19, 24.1% in ages 20–29, and 32.7% in those aged 30 years and older (18). Importantly, CF-related diabetes has been linked to higher mortality (19) and poorer cardiovascular outcomes (20).

To address these emerging health concerns, CF-specific nutrition guidelines have progressed, suggesting that priorities for many should shift from the high-fat approach (1), often met through energy-dense nutrient poor (EDNP) foods associated with cardiometabolic diseases (3), to achieving better diet quality in alignment with general population nutrition guidelines (1). The need for early counselling of individuals commencing VST treatment, with particular emphasis on balanced dietary choices and the importance of regular physical activity has been emphasised (17). However, research indicates that adults with CF continue to report poor diet quality (3, 21). A recent Irish study found half of adult participants with CF (*n* = 68) fell short of recommended increased energy targets despite having a high intake of fat and EDNP foods, as measured via both comparisons to national dietary guidelines and evaluation against a validated diet quality index (Healthy Eating Index – 2020) (21). Additionally, clinicians are grappling with how best to address the emerging issue of weight management in CF in the VST era and may lack specific weight management knowledge and skills (22).

Qualitative research allows for in-depth exploration of complex phenomena by capturing the nuanced experiences and perspectives of participants (23, 24). Previously qualitative studies related to diet with adults with CF have reported confusion on how to eat healthily for CF alongside co-morbidities like diabetes (25) and concerns in relation to weight gain, body image and dietary health implications (26). Qualitative studies have also revealed diet related experiences unique to this population such as not viewing food as a source of pleasure but as a treatment to endure to meet high energy targets (25).

In the context of significant health improvements resulting from VSTs, the clinical community faces the challenge of mitigating health risks associated with poor dietary habits and addressing the increasing prevalence of diet-related chronic diseases within the CF population. To effectively develop and evaluate personalised nutritional strategies for adults living with CF, further qualitative research on their dietary perceptions and experiences during the transition from traditional dietary recommendations to modulator era dietary guidelines and clinical practices is warranted. This study aimed to explore in depth the dietary perceptions and experiences of Irish adults living with CF in an era of widespread VST implementation.

## 2. Methods

### 2.1 Study design and population

This qualitative study was conducted and reported adhering to COREQ guidelines (Appendix 1) (COREQ, 27). Data were collected via online semi-structured interviews and focus groups. A demographic and self-reported health questionnaire administered via the Electronic Data Capture system (EDC), Castor, Version 1.6 (Ciwit B.V., The Netherlands), a virtual platform designed for handling clinical information, collected and stored data on demographic and clinical variables (e.g., age, gender, BMI, ling function, income status). The study protocol (28) was approved by the University of Limerick Institutional Review Board and relevant hospital ethics committees, with all participants providing written informed consent.

#### 2.1.1 Participant recruitment

Participants were recruited from a parallel quantitative study exploring dietary intakes and quality in adults living with CF (21). Recruitment for the quantitative parallel study was conducted through CF Ireland membership forums and CF centres/clinics across Ireland. Dietitians initially distributed recruitment emails via clinic mailing lists and actively promoted the study during clinic visits. In one centre with a high volume of PwCF, the dietitian pre-screened participants before inviting them via email or in person. In all other centres, eligibility was assessed using a self-report Microsoft Forms questionnaire developed for the study’s inclusion and exclusion criteria. From those who completed all components of the quantitative study (*N* = 73), participants were subsequently invited by email to participate in this qualitative study. Recruitment for the qualitative study took place from November 2021 to August 2023. The initial aim was to recruit approximately 30 adults living with CF using this convenience sampling method (29). Saturation point was ultimately used to assess sample size sufficiency. Saturation point can be described as information redundancy, or more specifically, when no new codes or themes are generated from data (30). Saturation was determined by two authors (CG & KR).

#### 2.1.2 Participant eligibility

*Inclusion criteria*: Adults (≥18 years old) with a diagnosis of CF, living in Ireland. *Exclusion criteria:* Receiving enteral nutrition, following a prescription diet for another medical condition (e.g., coeliac disease), were pregnant or have previously received an organ-transplant. Adults were required to be on a stable medical regimen for at least four weeks prior to commencing the study with no recent pulmonary exacerbations involving the administration of oral or intravenous antibiotics or glucocorticoids.

### 2.2 Data collection

#### 2.2.1 Semi-structured interviews and focus groups

Semi-structured interviews and focus groups with adults living with CF were conducted by a male PhD student trained in facilitation and qualitative research, with prior qualitative data collection and analysis experience and a primary qualification in nutrition science. The researcher had no prior relationship with participants. Questions and prompts were developed and piloted through consultation with a public and patient involvement (PPI) committee. The questions addressed three broad topics: views on diets advised for PwCF; current consumption patterns; enablers and barriers to eating a healthy diet. The readability of interview and focus group questions were measured using the Flesch-Kincaid readability test, a validated test taking into account readability level and education level in producing scores (31). After modifying the questions accordingly, an overall Flesch-Kincaid reading age of 5.9 (8-9^th^ grade: plain English) was obtained and a protocol was prepared (Appendix 2). Participants had the option to take part in either an online semi-structured interview or online focus group depending on their preference. Dates and times for interviews / focus groups were organised based on participant availability and conducted online via Microsoft Teams. Participants received a meeting link and set up instructions by email. Once eligibility was confirmed, participants were provided with a participant information and consent forms which could be completed online or in paper format. Written consent was obtained through the original consent forms completed at the start of the parallel quantitative study, offered in both paper and electronic formats, and verbal consent was reconfirmed at the beginning of the Microsoft Teams call. Interviews and focus groups were audio recorded through Microsoft Teams.

#### 2.2.2 Qualitative research assumptions

This study adopted a realist paradigm, acknowledging that while participant experiences reflect an external reality, the analysis process involves interpretation by the researcher and this process is inherently subjective. Ontologically, it assumes that participants’ accounts of their lived experience provide valid knowledge of their reality. Epistemologically, the study acknowledges the researcher’s role as an interpreter of these accounts, with their own perspectives influencing data analysis, consistent with Braun and Clarke’s reflexive thematic analysis approach (32).

### 2.3 Analysis

#### 2.3.1 Qualitative data

Audio recordings were automatically uploaded to Microsoft Streams after the completion of each interview or focus group. Recordings were converted to mp4 files and transcription was carried out via Microsoft Word. Mp4 files were entered into Microsoft Word via the transcribe function. Transcripts were cleaned and confidential information was removed prior to analysis. Transcripts were then imported into NVivo (QSR International Pty Ltd.), a qualitative analysis software, for analysis using an inductive thematic analysis method described by Braun and Clarke (33). This six-phase approach to thematic analysis entails: 1). data immersion; 2) generating codes; 3) generating themes; 4) reviewing potential themes; 5) defining and naming themes; 6) producing a report.

Initially, transcripts were read several times to immerse the researcher in the data, familiarise themselves with the participants use of language and descriptions and prepare further analysis (33). The first author coded all data. Through an inductive process, codes or labels identifying a feature of the data relevant to the research question were developed. For example, in the codebook, “time constraints” was used to label instances where participants described limited time as a barrier to healthy eating. After an initial transcript was coded, a co-author (KR) coded the same transcript independently and codes were compared and discussed. The remaining transcripts were coded by the first author with regular meetings with a co-author (KR) to refine the name and definition of codes and to reflexively consider researcher influence on the analysis process. Codes were reviewed and collated to develop subthemes and themes. A theme refers to a pattern of meaning that captures something significant in relation to the research question. In identifying themes, coherence, relevance to the research aims, support from multiple data points, recurrence across transcripts, and capacity to provide insight into participants’ experiences were assessed.

#### 2.3.2 Demographic and clinical data

SPSS® Statistics for Windows, Version 29 (IBM Corp., Released 2022) was used to complete statistical analysis of quantitative demographic and clinical data. The Kolmogorov Smirnov test assessed the normality of data distributions, where a significance of *p* >0.05 identified data as normally distributed. Based on this test, where appropriate, data was displayed as means ± standard deviations (SD), medians and interquartile ranges (IQR), and frequencies (*n* and %).

## 3. Results

### 3.1 Demographic and Clinical Characteristics

Of *N* = 73 participants who enrolled in the parallel quantitative study, *n* = 29 (mean age: 35.0 ± 10.1; female: 37.9%) participated in either an interview or focus group. The median BMI of the group was 24.8 (5.9) kg/m^2^, with a mean FEV_1_% of 78.2 ± 27.6% reported and 86.2% reported to be taking VSTs. A total of 20 PwCF took part in semi-structured interviews, and nine in focus groups. The overall demographic and clinical characteristics of those who participated in interviews and focus groups are displayed in Table 1. Participant-specific demographic and clinical characteristics are shown in Table 2.

**Table 1.** Demographic and clinical characteristics of Irish adults living with CF who participated in an interview or focus group.

|  | **$n = 29$** |
| --- | --- |
| Age (y) (range: 19-62) | $35.0 \pm 10.1$ |
| Female $n(\%)$ | 11 (37.9) |
| BMI ($\text{kg/m}^2$)* | 24.8 (5.9) |
| BMI Classification $n(\%)$~ | |
| Underweight ($<20$) | 1 (3.4) |
| Normal (20-24.9) | 14 (48.3) |
| Overweight (25-29.9) | 12 (41.4) |
| Obese ($\geq 30$) | 2 (6.9) |
| $\text{FEV}_1\%$ | $78.2 \pm 27.6$ |
| $\text{FEV}_1\%$ Categories $n(\%)$ | |
| $<40\%$ | 3 (10.3) |
| 40-69.99% | 8 (27.6) |
| 70-79.99% | 2 (6.9) |
| $\geq 80\%$ | 16 (55.2) |
| Hospital Admission |  |
| Since last admission (days)* | 794.0 (1069.5) |
| *Last admission length (days)\** | 14.0 (8.0) |
| *Admitted in last year n(%)* | 20 (69.0) |
| Highest Education n(%) |  |
| *Secondary level* | 3 (10.3) |
| *Third level‡* | 18 (62.1) |
| *Postgraduate level* | 8 (27.6) |
| Income Status n(%) |  |
| *Full income* | 16 (55.2) |
| *Part-time income* | 5 (17.2) |
| *No work-based income* | 8 (27.6) |
| Area of Living n(%) |  |
| *City* | 12 (41.4) |
| *Town* | 5 (17.2) |
| *Village* | 3 (10.3) |
| *Countryside* | 9 (31.0) |
| Living Situation n(%) |  |
| *Living alone* | 3 (10.3) |
| *Living with housemates* | 8 (27.6) |
| *Living with family/relatives* | 18 (62.1) |
| Clinical Characteristics n(%) |  |
| *Pancreatic insufficiency* | 24 (82.8) |
| *CF-related diabetes* | 9 (31.0) |
| *GORD* | 6 (20.7) |
| *CF-related liver disease* | 6 (20.7) |
| *CF-related bone disease* | 2 (6.9) |
| Medication / Supplement Use n(%)† |  |
| *VSTs* | 25 (86.2) |
| *Antibiotics* | 14 (48.3) |
| *Mucolytics* | 6 (20.7) |
| *Bronchodilators* | 11 (37.9) |
| *Insulin* | 9 (31.0) |
| *Steroids* | 3 (10.3) |
| *Fat-soluble vitamins* | 25 (86.2) |
| *Oral Nutritional Supplements* | 4 (13.8) |

Parametric values are presented as mean ± SD and non-parametric values as median (interquartile range).
Significance derived from a $p$ -value of $<0.05$ .
\*Non-parametric.
~World Health Organisation cut-off values (34).
†Chronic medications and supplements used on a regular basis.
‡University, college or vocational education.
Abbreviations: BMI, body mass index; FEV<sub>1</sub>%, percentage of predicted force expiratory volume for age, sex, ethnicity, and height; GORD, gastro-oesophageal reflux disease; VSTs, Variant-specific therapies SD, standard deviation.

**Table 2.** Participant-specific characteristics of adults living with CF who participated in an interview or focus group.

|  | Pseudonym | Age Category<br>(years) | Gender | BMI (kg/m<sup>2</sup>)* | Modulator Use | FEV<sub>1</sub>% |
| --- | --- | --- | --- | --- | --- | --- |
| P1 | Mary | 25-39 | female | Normal | yes | ≥80% |
| P2 | Paul | 40+ | male | Overweight | yes | 40-69.99% |
| P3 | Anne | 25-39 | female | Overweight | yes | 40-69.99% |
| P4 | Ciaran | 25-39 | male | Overweight | yes | ≥80% |
| P5 | Haleigh | 25-39 | female | Overweight | yes | ≥80% |
| P6 | Killian | 25-39 | male | Overweight | yes | 70-79.99% |
| P7 | James | 25-39 | male | Obese | yes | 70-79.99% |
| P8 | Sean | 25-39 | male | Normal | no | ≥80% |
| P9 | Aidan | 25-39 | male | Normal | yes | 40-69.99% |
| P10 | Ross | 40+ | male | Normal | yes | 40-69.99% |
| P11 | Darragh | 40+ | male | Overweight | no | <40% |
| P12 | Oisin | 40+ | male | Overweight | yes | ≥80% |
| P13 | Serena | 25-39 | female | Obese | yes | ≥80% |
| P14 | Stephen | 25-39 | male | Overweight | yes | ≥80% |
| P15 | Peter | 25-39 | male | Normal | yes | 40-69.99% |
| P16 | Mark | 25-39 | male | Normal | yes | ≥80% |
| P17 | Neil | 40+ | male | Overweight | yes | ≥80% |
| P18 | Vivienne | 18-24 | female | Normal | no | ≥80% |
| P19 | Fiona | 25-39 | female | Normal | yes | <40% |
| P20 | Padraig | 40+ | male | Overweight | yes | 40-69.99% |
| P21 | Bill | 25-39 | male | Overweight | yes | ≥80% |
| P22 | Jackie | 18-24 | female | Normal | yes | ≥80% |
| P23 | Nathan | 25-39 | male | Normal | no | ≥80% |
| P24 | Roisin | 40+ | female | Underweight | yes | <40% |
| P25 | Emma | 25-39 | female | Normal | yes | 40-69.99% |
| P26 | Emily | 25-39 | female | Normal | yes | ≥80% |
| P27 | Sophie | 25-39 | female | Normal | yes | 40-69.99% |
| P28 | Joe | 18-24 | male | Normal | yes | ≥80% |
| P29 | Eric | 40+ | male | Overweight | yes | ≥80% |

\*World Health Organisation cut-off values (45): Underweight (<20 kg/m<sup>2</sup>); Normal (20-24.9 kg/m<sup>2</sup>); Overweight (25-29.9 kg/m<sup>2</sup>); Obese (≥30 kg/m<sup>2</sup>).
Abbreviations: BMI body mass index, FEV<sub>1</sub>%, percentage of predicted force expiratory volume for age, sex, ethnicity, and height.

### 3.2 Qualitative Themes and Subthemes

Five themes were generated from the data: (1) Legacy effects of historic CF diet (2) Evolving relationship with food, (3) Weight gain and altered body image (4) Reclaiming authority over the body, (5) Navigating dietary choices. An overview of themes and corresponding subthemes is provided in Appendix 3.

#### 3.2.1 Theme 1: Legacy effects of historic CF diet

Most participants identified that clinical dietary advice and interventions they received in childhood continue to impact their adult lives, shaping their food preferences and perceptions of what constitutes a healthy diet. While some acknowledged healthy eating recommendations, others held deeply ingrained beliefs about maximising calories and fat.

> “*I think definitely you still need your calories and your fat… I think if I even went off it for a week, I probably notice that I would probably be losing weight*.” Fiona (F, Age 25-39)

Many described a childhood diet that focused primarily on increasing caloric intake, often neglecting the quality of those calories. As one participant noted, “*I’ve always grown up where calories were always pushed on me no matter what kind of calories you need, even if they’re empty calories*.” Padraig (M, Age 40+)

This legacy of prioritising energy density over diet quality or balance contributed to early eating habits that were misaligned with appetite and a preference for high-fat foods. As Ciaran (M, Age 25-39) expressed “*… at one stage, desperation… I was getting maybe three takeaways a week.”*.

Some described how childhood experiences of clinical interventions such as feeding tubes or feedback from clinicians regarding their weight instilled a persistent anxiety about weight stability and medical scrutiny of their weight in adulthood. In the following quote Anne (F, Age 25-39) expresses a desire to lose weight while fearing repercussions if she does so.

> “*I’m also afraid that if I lose weight, they’ll be like, ‘see, you shouldn’t have got your feeding tube out’… even though I’m not really happy with the weight I am*.”

Some described weight-focused self-monitoring, with varying fears of clinician responses to them being underweight or overweight.

> “*What I’d be conscious of now is I know I have an appointment next week and I’m thinking in my head… if the weight has either stayed the same or gone down… they’re going to turn to me and say like ‘you need to put on more weight’*.” Mary (F, Age 25-39)

#### 3.2.2 Theme 2: Evolving relationship with food

Many participants articulated that their relationship with food has been reshaped since the introduction of CFTR modulators. Participants consistently described a sense of liberation and transformative change following the introduction of modulators. “*Once I started the modulators… I actually felt like I gained a superpower… is this how normal people feel? Their food just digests, like, immediately.*” Haleigh (F, Age 25-39)

For many, food transitioned from a burdensome medical requirement and a “*chore*” to a source of pleasure, normalcy, and even excitement where they could “*enjoy food for the first time in a long time”* Mark (M, Age 25-39)

> “*Back then, I ate because I had to. But it was minimal. Now, you can’t stop me eating. I just love everything.*” Sophie (F, Age 25-39)

Many participants articulated how modulator treatment reduced the constant pressure to eat high-fat, high-calorie meals and allowed for a more intuitive approach to eating. As described by Fiona (F, Age 25-39):

> “*One time I’d be constantly thinking I have to eat. Whereas now I can do the whole normal thing… Kaftrio eased all the pressure off you. Once you had to keep eating… Kaftrio held your calories.*”

A new sense of normalcy coupled with improved energy levels in some participants after initiating modulator therapy prompted a wider re-engagement with health and lifestyle, in some cases prompting reassessment of routines and adoption of healthier habits related to diet and exercise. One participant explained: “*I started taking an interest in exercise… that all amalgamated into me just wanting to take a better interest in my diet.*” Jackie (F, Age 18-24)

For some, these changes were sustained and integrated into everyday life, whereas for others they were more tentative or short-lived, reflecting ongoing negotiation between old habits, new possibilities, and fluctuating health.

> “*I was trying to watch what I ate. I did it for one week and I was weighed out my food, but I got fed up. I got sick of doing that.*” Anne (F, Age 25-39)

#### 3.2.3 Theme 3: Weight gain and altered body image

Many participants described how modulator therapies and other bodily changes have transformed their weight, body image and feeling of visibility. While some embraced weight changes as markers of normalcy or restored health, others struggled with fluctuating appearance and perceived societal expectations. Amongst participants who were now taking VSTs, the majority reported weight gain since commencing modulator therapy and for many, this weight gain and dietary changes represented something closer to “*normal life*”. Several participants expressed joy or relief at being able to eat like their peers, feeling less restricted and defined by their dietary needs. However, for many the bodily changes that accompanied modulator therapy were experienced as rapid and unpredictable including sudden weight gain, altered appetite, a change in how their bodies metabolised food and perceived changes in body composition.

> “*I gained weight in ways that I never had before… it didn’t feel like natural weight gain or fat stores… it almost felt like inflammation*.” Haleigh (F, Age 25-39)

Some participants attributed difficulty with weight management to the drugs themselves, leading a few to modify or discontinue treatment:

> “*My weight ballooned up… I felt a lot of other health issues were linked to that CF drug.*” James (M, Age 25-39)

Challenges losing weight were reported by some, including Ciaran (M, Age 25-39) who described no weight change even after completing a marathon.

> “*I did a marathon in April… and I didn’t lose a kilo… that will tell you how hard it’s after getting to lose any weight*.”

The flux of weight, appetite, and digestion left many navigating unfamiliar circumstances, sometimes without support or sufficient information from clinicians.

> “*When I was starting Trikafta, no one was going to say your sugar cravings or salt cravings are going to diminish… just learn as you go.*” Stephen (M, Age 25-39)

While some described weight gain in exclusively positive terms, many described a negative shift in their body image, moving from a longstanding anxiety about being underweight, to more recent challenges related to weight gain and perceptions of being overweight:

> “*Once it does happen, it happens really fast that it can be a bit of a shock.*” Vivienne (F, Age 18-24)

Weight gain, previously a clinical goal, now prompted internal conflict and renewed self-consciousness, particularly among women who felt pressure to conform to broader societal ideals.

> “*…you’re competing with gender expectations around ‘when you’re a woman you should be tiny’*.” Mary (F, Age 25-39)

Unsolicited comments related to weight from peers and healthcare professionals were frequently reported, heightening participants’ body awareness and making them feel scrutinised or uncomfortable.

> “*I suddenly had a good bit of weight on and people were commenting… it still gets me down, I would still struggle with it to this point.*” Padraig (M, Age 40+)

Bloating, steroid-related changes, or visible differences associated with CF added to this sense of scrutiny, creating tension between how many participants felt and how they believed others perceived them.

> “*I would often not be in love with the physique I’d see… bloated and gassy… you kind of feel like you’ve a gut on you*.” Sean (M, Age 25-39)

#### 3.2.4 Theme 4: Reclaiming authority over the body

In navigating diet and nutrition in the context of CF management, participants describe the need to readjust where professional knowledge ends, and personal authority begins.

While many participants reflected on having a positive relationship with the multidisciplinary team, especially when spoken to in an open and collaborative way, several participants described strained or conflicted relationships with clinicians around diet, particularly when advice from clinicians felt outdated, overly prescriptive, or dismissive of personal experience:

> “*We were just at war*” Sophie (F, Age 25-39)

Experiences varied and frustration was directed not only at dietary guidance but also at the communication styles used. While some recounted positive experiences with clinicians who were responsive and engaged, others felt unheard or constrained:

> “*They don’t listen to you, so you have to own your own health.*” Ross (M, Age 40+)

Others expressed a desire for more openness from healthcare professionals in acknowledging diverse diets, experiences, and preferences:

> “*If the dietitian was more open to different ways of doing things…”* Anne (F, Age 25-39)

Many participants described exercising authority and control over food choices and eating behaviours, monitoring and adjusting their intake, and sourcing information independent from their healthcare providers.

> “*I would look at it most days through MyFitnessPal to keep an account of what I’m eating and my macros*.” Darragh (M, Age 40+)

Digital platforms, social media groups, and personal research were frequently cited as alternative sources of knowledge:

> “*I will have an idea like ‘Oh, should I eat this kind of food’ and I’ll go off and Google it… from a few different sources*.” Mary (F, Age 25-39)

Several participants articulated the emotional and psychological burdens of chronic illness, emphasising the need for *“…a more holistic approach…*” Sophie (F, Age 25-39) and calling for person-centred, collaborative approaches from clinicians in CF care.

> “*You’re treated as a number. You’re treated as a condition. You’re not treated as a human being.*” Padraig (M, Age 40+)

Participants expressed a strong desire for deeper connection, “*You need to have that close relationship with the team…*” Stephen (M, Age 25-39), empathy, and recognition from healthcare providers, relationships built on mutual respect, trust, shared decision making and personalised care rather than standardised protocols:

> “*So I think really to just have a dietitian… willing to try new things and to listen to my experiences*.” Haleigh (F, Age 25-39)

Strategies and clinician communication that was patient-centred and acknowledged lived experience was praised:

> “*I’ve always appreciated that open line of communication… they’ve always been very supportive (of choice to be vegetarian).*” Jackie (F, Age 18-24)

When professionals engaged openly and without judgement, it made a lasting impact:

> “*I just remember [names respiratory clinician] listened to me. I was impressed by it*.” Ross (M, Age 40+)

#### 3.2.5 Theme 5: Navigating dietary choices

Most participants acknowledged that diet plays a crucial role in managing CF. However, their dietary choices were also influenced by a combination of practical factors such as time, budget, and accessibility, and individual factors such as mood, comfort, and the impact of early dietary guidance.

The majority of participants’ accounts illustrated how eating practices were shaped not only by beliefs about the correct diet and physical needs but also by emotional states, life history, and the constraints of daily routines. Food was frequently described as a source of comfort, pleasure, or escape, especially in the context of chronic illness:

> “*Food is my vice, it’s a comfort thing… I have a multitude of different medical ailments, so food is a bit of an escape*.” Padraig (M, Age 40+)

For some, shifts in health status directly influenced appetite and cravings. Periods of ill-health often led to a preference for “*high-fat and sugar*” snacks. Day-to-day eating was also shaped by mood, motivation, and life stages:

> “*I’ll veer towards healthier eating when I’m training…It changes with my mood*.” Serena (F, Age 25-39)

Practical barriers such as fatigue, and convenience played a major role in shaping food choices, yet time pressure was almost unanimously highlighted as a major barrier for healthier eating. Participants often relied on quick, easy-to-prepare meals:

> “*After a long day… the last thing I want to do is cook.*” James (M, Age 25-39)

While some framed eating patterns as responses to daily pressures, others acknowledged that discipline and motivation played a role:

> “*It’s just about the determination and commitment… the discipline probably needs to be stronger for me to follow a healthier diet*.” Neil (M, Age 40+)

For some participants, finances shaped the variety of their diets. While not always the dominant barrier, cost influenced the ability to buy preferred foods, try new options, or maintain a consistently nutrient-rich diet.

> “*Strawberries are expensive. Everyone knows that…I couldn’t afford a lot of the nicer foods like fruit, veg, nuts…the bare minimum*” Jackie (F, Age 18-24)

Access to food and cooking facilities varied across life stages and contexts, influencing participants’ autonomy in meal preparation. For some, adulthood brought greater control and variety:

> “*Now I have more access to kitchens and cooking facilities… I often cook extra dinner and use the leftovers for lunch*.” Sean (M, Age 25-39)

Workplace location and set-up also influenced eating patterns. Participants valued arrangements that allowed more control over timing and type of food:

> “*Working from home is great from a CF perspective… you have control over your meals and snacks*.” Oisin (M, Age 40+)

## 4. Discussion

This study aimed to explore diet related perceptions and experiences amongst adults living with CF in the context of widespread implementation of VSTs. To the authors knowledge, the study is the first to provide a detailed qualitative exploration of how Irish adults living with CF perceive and experience diet in the modulator era. Five interconnected themes were identified: *Legacy effects of CF historic diet*; *Evolving relationship with food*; *Weight gain and altered body image*; *Reclaiming authority over the body*; *Navigating dietary choices*. The themes reveal how historic dietary practices, treatment transitions, and broader psychosocial contexts combine to shape orientations towards diet and health in the modulator era.

### 4.1 Dietary influences and transition

Across themes, multiple factors were reported that influence dietary choice and behaviour in adults with CF. Many of these factors have been extensively reported previously in studies with other populations such as emotional and mood related influences on eating, time constraints driving reliance on convenience foods, financial limitations affecting food variety and beliefs about nutrition (35). Additionally, we found that, among adults living with CF the childhood dietary advice and interventions they experienced that were focused on high-fat, high-calorie intakes had an enduring impact into adulthood. This impact continued despite the widespread use of VSTs. Participants described these childhood experiences as influencing their long-standing preferences for EDNP foods and anxiety about weight and clinical scrutiny into adulthood. Enduring beliefs about the need for EDNP foods were articulated by some participants. Previous research has identified this reliance on EDNP food in children (*n* = 80) (36) and adults living with CF (3). In the modulator era, this pattern appears to persist, with a cross-sectional study of adults living with CF (*n* = 68) reporting continued reliance on EDNP foods. Despite this, only half of participants achieved CF-specific energy targets (110–200% of the estimated average requirement) (21). A retrospective cohort study (*n* = 40) also reported that after commencement of ETI, participants still maintained a high EDNP food intake despite reducing caloric intake (37). Insights compliment earlier research (*n* = 10) which reported childhood experiences as formative in shaping dietary behaviours among PwCF (26). However, unlike earlier studies, these findings point to a new tension, wherein people with CF must reconcile the health gains from modulators with nutritional guidance that was historically centred on survival rather than long-term wellbeing. This creates a challenging dietary transition, in which participants are simultaneously unlearning entrenched habits shaped by past survival-focused advice and adapting to altered physiological needs.

Participants described a shift from diet being a “*chore*”, to greater pleasure and autonomy in eating since starting modulator therapy. This aligns with previous qualitative research in the UK (*n* = 20) where most participants reported no pleasure in food, but that food was another treatment to endure, due to the need to achieve high energy targets (25). However, the current study extends these insights by highlighting how this relationship with food is evolving in the modulator era, with some participants beginning to experience a new sense of enjoyment and control over their eating. As CF-specific dietary recommendations shift from a focus on survival to an emphasis on wellbeing and diet quality (1), the burden of trying to achieve high energy targets may begin to be mitigated for many PwCF. Many participants also now articulate food as a source of normalcy and empowerment. This mirrors findings from other chronic conditions undergoing therapeutic revolutions, such as rheumatoid arthritis (38), where improved treatment outcomes created opportunities to redefine their identity and alter their lifestyle (39, 40). However, the shift in CF was not described as linear. For some, modulators reduced pressure and allowed for some to adopt intuitive eating. In others, weight gain, changing appetites, or co-morbidities introduced new dietary burdens. These findings show that the shift to ‘normal’ from modulators is not just about physical health, but also about how people see themselves, how others see them, and their everyday experiences. Given the wide range of experiences, the findings highlight the importance of individualised and holistic approaches in clinical practice to support PwCF.

Participants’ accounts also show that food choices are closely tied to the wider circumstances of daily life. Fatigue, limited time, financial pressures, and emotional wellbeing all combined with long-standing dietary habits to influence how participants ate. These findings are consistent with CF research (41) and studies of chronic illness more generally (42, 43), which highlight how social and structural factors shape choices like dietary behaviour. This suggests that effective interventions need to go beyond individual motivation, addressing the practical and social realities that affect food access, preparation, and consumption.

### 4.2 Body image and visibility in the modulator era

While historic body image concerns were linked to being underweight, many participants articulated that modulators have introduced new anxieties related to weight gain, body composition, and heightened social visibility. A critical review on BI in CF published in 2012 reported that females had better body image compared to males and other healthy peers, with findings marked as a sequential outcome of being a lower weight, common amongst PwCF (44), which fits sociocultural norms of attractiveness for females (45). Differing from past research, in the current study, female participants voiced more concerns related to their body image, with increases in BMI and weight related to modulator therapies contributing to these concerns, alongside expectations placed upon women in relation to their appearance. Nonetheless, it must be noted that body image concerns were not exclusive to female participants. Previous qualitative studies amongst young people and adults living with CF identified body image as an important and concerning topic for PwCF (26, 41, 46). Participants in the current study frequently reported feeling unheard by clinicians and pressured to gain weight which sometimes misaligned with personal desires. This aligns with findings by Helms *et al.* (46) who also noted that participants were reluctant to raise body image concerns with clinicians, preferring healthcare providers to initiate these conversations as part of routine care to make patients feel more comfortable discussing the topic. By shifting focus from weight gain pressures to initiating conversations about body image, this could potentially foster a greater sense of professional support and encourage more collaborative patient-clinician relationships. Moreover, Helms *et al.* (46) (*n* = 20) reported that 85% of CF participants had never had a conversation with a health care provider about their BI or how they felt about their bodies, further highlighting the importance of initiating these conversations.

### 4.3 Movement towards collaborative care

Another central finding was the reclaiming of authority over dietary decisions. Some participants described tension between professional dietary advice, often perceived as outdated or overly prescriptive, and their own knowledge and experiences. This builds on a growing body of literature advocating for patient-led, participatory models of care in CF, other chronic illnesses and in general patient care (47–49). Importantly, resistance was not framed as outright rejection of medical expertise but as a desire for more holistic, empathetic, and collaborative interactions with dietitians and the multidisciplinary team. Participants valued healthcare providers who engaged in shared decision-making, respected individual dietary preferences, and acknowledged the psychological dimensions of food. This aligns with qualitative CF research in the past where participants described that positive rapport with healthcare providers would enhance openness in discussing sensitive topics (41, 46). At the same time, some participants described feeling “*treated as a number*” and not as a “*human being’*’, echoing past findings that clinicians were perceived as prioritising weight gain over overall wellbeing. In particular, threats of feeding tube placement reinforced the belief that clinicians cared more about weight than the person (46). This concern persisted in the current study, with some participants who had past experience of feeding tubes expressing anxiety about reinsertion if they did not continue to follow weight gain advice. This suggests that evolving CF nutritional care must move beyond calorie-based metrics to embrace a broader conception of health, identity, and patient autonomy.

### 4.4 Implications for practice

Collectively, these findings reinforce the ongoing shift in nutritional care in the modulator era, moving from a survival focus toward supporting overall health and well-being (1). Clinical teams must move beyond survival-focused, high-fat paradigms to support PwCF in developing nutrient-dense dietary patterns that align with general population health guidance while remaining sensitive to individual histories and embodied experiences. Dietitians may require additional training in motivational interviewing, collaborative care, and body image counselling to effectively support patients through this nutritional transition (22). Importantly, clinical guidelines should acknowledge the psychosocial challenges of shifting dietary norms, rather than assuming a straightforward adoption of new recommendations. Further research is needed to examine how dietary transitions unfold longitudinally across different stages of life and disease progression, and how healthcare teams can best support this process. Intervention studies co-designed with PwCF are warranted to develop practical, patient-centred approaches to diet that balance clinical recommendations with lived experiences.

### 4.5 Strengths and limitations

This study is strengthened by its large qualitative sample relative to prior CF studies, the use of both interviews and focus groups, and its grounding in PPI during study design. The inclusion of participants across a range of ages, disease severities, and treatment experiences enhances the richness and transferability of findings. Nonetheless, online data collection may also have limited rapport or excluded those with digital access barriers. Another limitation is that females comprised only 37.9% of the sample, which may have limited the breadth of female perspectives captured, particularly given that body image and dietary experiences can differ by gender. While thematic analysis was conducted rigorously and reflexively, interpretation remains situated within the researchers’ perspectives. Finally, although saturation point is often considered a gold standard approach for sample size sufficiency in qualitative research, Braun and Clarke (50) have recently argued that it may not always be the most appropriate criterion, particularly in inductive approaches. In such designs, insights from earlier interviews can shape the focus of subsequent ones, making the point of complete information redundancy difficult to establish. Nonetheless, in this study, the authors felt that saturation was achieved, as no new codes emerged during the final interviews.

## 5. Conclusion

This study provides the first in depth qualitative exploration of how Irish adults living with CF perceive and experience diet in the modulator era. By identifying five interconnected themes: *Legacy effects of historic CF diet*; *Evolving relationship with food*; *Weight gain and altered body image*; *Reclaiming authority over the body*; *Navigating dietary choices*; the findings highlight the complex ways in which historical practices, treatment transitions, and everyday contexts shape dietary orientations.

A central insight is that early nutritional strategies, while lifesaving, continue to influence food preferences, anxieties, and clinical interactions into adulthood. Modulator therapies have begun to transform these dynamics, creating opportunities for greater enjoyment and autonomy in eating. Yet, this transition is not linear. For some, modulators alleviate burdens and enable intuitive eating, while for others, new challenges such as weight gain and shifting appetites emerge. These experiences underscore that dietary change cannot be addressed solely through clinical targets, but must engage with how people understand their health, bodies, and identities.

Importantly, participants described reclaiming a sense of agency over dietary decisions and sought more collaborative, empathetic relationships with clinicians. Food was framed not only as fuel, but as a source of empowerment, normalcy, and social belonging. However, barriers such as fatigue, time, finances, and emotional wellbeing continue to constrain choices, highlighting the need for interventions that address both psychosocial and structural realities.

Taken together, these findings suggest that nutritional care in the modulator era must move beyond a survival focus and should embrace holistic, patient-centred approaches that balance medical guidance with lived experiences. Supporting adults with CF in this transition will require further re-evaluation of dietary guidelines, enhancing clinical training in collaborative care, and co-designing interventions with PwCF to ensure dietary recommendations are practical, empowering, and sustainable.

## Abbreviations

CF: Cystic Fibrosis
CFTR: Cystic Fibrosis transmembrane conductance regulator
PwCF: people living with Cystic Fibrosis
VST: variant specific therapies
BMI: body mass index
EDNP: energy-dense nutrient poor
EDC: Electronic Data Capture system
PPI: public and patient involvement.

## 6. Declarations

### Ethical approval

The study was conducted in accordance with the Declaration of Helsinki and was approved by the University of Limerick Education and Health Sciences, Research and Ethics Committee, University Hospital Limerick (Ref: 090/2021), and Cork University Hospital (Ref: ECM 4 (o) 11/01/2022 & ECM 3 (bb) 22/02/2022), University Hospital Galway (Ref: C.A. 2709), and St. Vincent’s University Hospital (Ref: RS22-022) Research and Ethics Committees for all aspects of the study. All participants provided written informed consent prior to study enrolment.

### Data availability statement

The data underlying this study consist of qualitative in-depth interview transcripts containing personal information about adults living with cystic fibrosis, as well as other contextual details that, although not individually identifiable, could potentially identify participants when combined, given the small size of the adult cystic fibrosis community in Ireland.

In accordance with the ethical approval granted by the University of Limerick and Cystic Fibrosis Hospitals/ Clinics, and the conditions outlined in participants’ informed consent, full interview transcripts cannot be shared publicly to protect participant confidentiality.

However, a comprehensive set of supporting materials is provided with this submission to ensure transparency and reproducibility. These include the COREQ reporting checklist, interview topic guide, thematic framework (themes and subthemes table), and a detailed codebook containing number of references, and number of sources coded. De-identified excerpts supporting each theme are presented within the manuscript. Additional de-identified information (such as the analytic structure linking codes to subthemes and themes) is available in the supplementary files. Those seeking access to further anonymised data may contact the corresponding author, Cian Greaney (School of Allied Health, University of Limerick), subject to approval from the Ethics Committees and a data-sharing agreement.

## Acknowledgements

The authors would like to acknowledge the valuable contribution of the Cystic Fibrosis Public and Patient Involvement Group, Cystic Fibrosis Ireland, and the people living with Cystic Fibrosis who took part in the study.

## Author contributions

Cian Greaney: Conceptualization, Methodology, Validation, Formal analysis, Investigation, Resources, Data curation, Writing – original draft, Writing – review & editing, Visualization, Project administration, Funding acquisition.

Katie Bohan: Investigation.

Sarah Tecklenborg: Resources, Writing – review & editing, Funding acquisition.

Ciara Howlett: Resources, Writing – review & editing.

Karen Cronin: Resources, Writing – review & editing.

Clodagh Landers: Resources, Writing – review & editing.

Mary Connolly: Resources, Writing – review & editing.

Derbhla O’Sullivan: Resources, Writing – review & editing.

Audrey Tierney: Conceptualization, Methodology, Investigation, Resources, Writing - review & editing, Supervision, Project administration, Funding acquisition.

Katie Robinson: Conceptualization, Validation, Writing – review & editing, Supervision, Funding acquisition.

